# Identification of severe cases with routine Pulse Oximetry use into the Integrated Management of Childhood Illness at Primary Health Centres level in West Africa: A cross-sectional study within the AIRE project in Burkina Faso, Guinea, Mali and Niger, 2021 - 2022

**DOI:** 10.1101/2024.10.14.24315439

**Authors:** Hedible Gildas Boris, Sawadogo Abdoul Guaniyi, Zair Zineb, Kargougou G Désiré, Agbeci Honorat, Méda Bertrand, Peters-Bokol Lucie, Jacques S Kolié, Louart Sarah, Ouédraogo Yugbaré Solange, Diakite Abdoul Aziz, Diallo Ibrahima Sory, Abarry Souleymane Hannatou, Neboua Désiré, Vignon Marine, Busière Sandrine, Lamontagne Franck, Ridde Valéry, Leroy Valériane, AIRE Research Study Group

## Abstract

**Background:** The Integrated Management of Childhood Illness (IMCI) guidelines is a symptom-based algorithm guiding healthcare workers in resource-limited countries to identify critically ill children under-5 in primary healthcare centre (PHC). Hypoxemia, a life-threatening event, is often clinically underdiagnosed. The AIRE project has implemented the routine use of Pulse Oximeter (PO) in IMCI consultations to improve the diagnosis of severe hypoxemia (SpO_2_ < 90%) and the management of severe cases at PHC level in Burkina Faso, Guinea, Mali and Niger. We measured the prevalence of IMCI+PO severe cases, and their associated social and structural factors among IMCI outpatients.

**Methods:** In 16 AIRE research PHC (4/country), all the children under-5 attending IMCI consultations, except those aged 2-59 months classified as simple case without cough or breathing difficulties, were eligible for PO use and enrolled in a cross-sectional study with parental consent. Severe IMCI+PO cases were defined as IMCI severe cases or those with severe hypoxemia.

**Results:** From June 2021 to June 2022, 968 neonates (0-59 days) and 14,868 children (2-59 months) were included. Prevalence of severe IMCI+PO cases was heterogeneous between countries: 5.0% in Burkina Faso, 6.1% in Niger, 18.9% in Mali and 44.6% in Guinea. Among neonates, 21.9% (95%CI: 19.3-24.6) were classified as IMCI+PO severe cases versus 12.0% (95%CI: 11.4-12.5) in older children. Severe hypoxemia was identified in 3.3% of neonates versus 0.8% in older children (p<0.001). The adjusted social and structural factors associated with disease severity commons to all four countries were: age <2 months or >2 years, IMCI-consultation delay >2 days, home to PHC travel time >30 minutes.

**Conclusion:** Despite between-country heterogeneity, the prevalence of seriously ill children under-5 including severe hypoxemia was high, particularly in neonates, and those without accessibility to PHC. Improving earlier access to primary healthcare and management of severe cases remains needed in West Africa.

- **What is already known on this topic**

- A few studies have reported the prevalence and correlates of severe illnesses with the IMCI guidelines using routine integration of pulse oximetry (PO) among all children under-5 at primary healthcare Centres (PHC) level in low-middle income countries (LMICs), and to our knowledge none has been done in West Africa.
- **What this study adds** This study:

- reports a high prevalence of severe cases using IMCI including PO among outpatient children under-5 attending IMCI consultation, and heterogeneous between countries (Burkina Faso, Guinea, Mali, Niger)
- shows that overall prevalence of severe cases was significantly twice higher in neonates (21.9%) than in children aged 2-59 months (12.0%). Similarly, the prevalence of severe hypoxemia was higher in neonates (3.3%) than in older children (0.8%).
- highlights the accessibility challenges to primary healthcare for children with serious illnesses, and inadequate decision about their specific care management.

- **How this study might affect research, practice or policy**

- This study provides original and reliable estimates for policy-makers to invest in earlier access to primary healthcare and better referral decision of severe IMCI cases to improve child health in West Africa.
- These indicators will be useful in assessing the added-value of PO integration into IMCI in LMICs and support scaling-up of PO into both national and international IMCI guidelines.

## Introduction

The Sustainable Development Goal 3 aims to reduce the under-5 mortality rate to lower than 25 deaths per 1000 live births by 2030 (1–3), but mortality among children under-5 still remains high: globally, 5 millions of children under-5 years of age died in 2021. Sub-Saharan African and Asian countries contribute highly to this mortality estimated at 74 deaths per 1000 live births in 2021 (4). West African countries are particularly concerned. There, child mortality is associated with fragile health systems (5), unskilled health care workers (HCW), and inaccurate diagnoses of severe diseases at primary healthcare centres (PHC) due to the lack of tools (6–9). In 1996, the World Health Organisation (WHO) has therefore proposed the Integrated Management of Childhood Illness (IMCI) Guidelines (10), revised several times (11,12) using a symptom-based algorithm guiding HCW to better identify and manage severe cases at the PHC level in resource-limited countries. These guidelines were adopted and adapted by each country to its own context. Some countries use a paper format, others an electronic one, and this is coupled with other strategies such as malaria control (including the use of bed nets impregnated with long-acting insecticide (13,14), malaria Rapid Diagnosis Tests (mRDT) in febrile children (15,16), and seasonal malaria chemoprevention (17) etc.), and to improve the management of malnutrition (18,19), digestive parasitosis, and other health programs targeting children under-5. However, despite the use of these IMCI guidelines, the diagnosis of severe illnesses remains difficult, inaccurate and delayed, leading to their inappropriate care management (20,21). Hypoxemia is therefore not well identified clinically by IMCI. Severe hypoxemia defined as low levels of oxygen in the blood (pulse blood oxygen saturation, Sp0_2_<90%) (22) is a common manifestation of severe illness, but very difficult to diagnose clinically (23,24). The prevalence of severe hypoxemia was estimated to 22% of sick inpatient neonates in Nigeria (23), 1.3% in outpatient in Uganda (25) and could reach 23% of children under-5 managed as outpatients at frontline in sub-Saharan Africa (26). Severe hypoxemia, a strong predictor of mortality should be better managed at the PHC level using pulse oximeters (PO) to enable its early detection with subsequent hospital referral for oxygen therapy (27).

The AIRE project (Amélioration de l’Identification des détresses Respiratoires de l’Enfant) has been implemented with a consortium of three NGOs (ALIMA, SOLTHIS, Terre des hommes) and the French Institute of Health and Medical Research (Inserm). It aimed to improve the detection of severe hypoxemia in children under-5 years of age, and their care management at PHC level by introducing the routine use of the PO integrated into the IMCI consultations in 202 PHCs in four West African countries: Burkina Faso, Guinea, Mali, Niger(28). A research operational study assessed this intervention. This cross-sectional analysis is aimed to estimate the prevalence of severe cases among under-5 outpatients at the PHC level when using IMCI guidelines integrating routine PO use. It seeks to examine their characteristics and the factors associated with severe cases.

## Methods

### Study sites

The AIRE operational research study took place in two health districts per country, in a total of eight district hospitals and 202 PHCs, including 16 research PHCs (four per country) where a specific operational research study has been conducted according to the published protocol (28). A baseline site assessment described the characteristics of the AIRE study context (29).

### Study design

A population-based cross-sectional study was conducted among IMCI children under-5 years of age managed in the 16 research PHCs, providing individual data to assess outcomes of PO introduction into IMCI consultations.

### Study population and inclusion criteria

From 2021, 14^th^ June to 2022, 20^th^ June, all the neonates (defined from 0 to 59 days) and children (2 to 59 months) attending IMCI consultations in the 16 research PHCs were screened by the site HCW using the national IMCI algorithms to classify and manage them based on their disease severity into three groups: green for simple cases (going back to home), yellow for moderate cases (observed and treated at PHCs then at home) and red for severe cases requiring urgent hospital transfer. The AIRE research study proposed that after each IMCI consultation, PO should be used according to child’ age and initial IMCI classification. PO use was implemented routinely into IMCI guidelines (based either on electronic or paper support) for all under-5 children attending IMCI consultations, except those aged 2-59 months classified as non-respiratory (without cough or breathing difficulties) simple cases. All the children initially classified as non-severe cases (green and yellow cases) using IMCI who had severe hypoxemia (SpO_2_< 90%) using PO joined the severe case (red) group. Then, the dedicated AIRE study team proposed the child inclusion to all those eligible for PO use. Those whose parents gave the written consent were included.

### Procedures

The procedures for the IMCI consultation and for carrying out mRDT had not been modified by the AIRE project. All clinical parameters (axillary temperature, weight, height…) were measured using thermometer, child scale, height scale…) in the triage room if available, or at the beginning of IMCI consultation by clinician. Then, PO (Acare Technology, Taiwan; AH-M1 S0002033) was used by onsite clinicians after IMCI classification to measured oxygen saturation in blood (SpO_2_) with appropriate probe according to age, and heart rate and reported these 2 parameters into register of consultation (paper-based or electronic) depending on what IMCI support was in force at the PHC.

Clinicians had received refresher training in IMCI but had not been trained in research procedures except a general briefing about Good Clinical Practice. A separate team was dedicated to research data collection. Data had been extracted by AIRE data collectors who were in general nurses, from paper-based register of consultation or from electronic data base set up by NGO Tdh in Burkina Faso and Markala health district in Mali.

### Data collection and definitions

Individual data collection had been carried out over the whole period of inclusion via electronic case report form with REDCap® software. These include socio-demographic data, clinical data about IMCI consultation including IMCI+PO classification and decision of care management.

In this work, neonates were defined between until 59 days of age, in conformity with the IMCI definition. Severe hypoxemia was defined as SpO_2_ level less than 90% and moderate hypoxemia when SpO_2_ is from 90% to 93% (25,30). Normal heart rate (or cardiac frequency) is defined as heart rate between 100 and 160 per minute in children aged from 0 to 1 year and from 90 to 150 for children between 1 and 3 years and from 80 to 140 for those aged between 3 to 5 years. Fever was defined as body temperature equal to or greater than 38°C using digital thermometer(31). Respiratory rate was only measured in children aged between 2 and 59 months according to IMCI guidelines. Fast breathing was defined as a respiratory rate greater than or equal to 50 breaths per minute for children aged from 2 to 11 months inclusive and greater than or equal to 40 breaths per minute for those aged from 12 to 59 months inclusive (12). Respiratory case was defined when the clinician identified at least one respiratory symptom such as coughing, fast or difficulty breathing, stridor …

### Statistical analysis

We first described the socio-demographics and clinical characteristics of children under-5 eligible for PO use during IMCI consultations, overall and by country: distribution by age, sex, number of people living in household, education level of household manager, income-generating activity of accompanying person, distance from home to PHC, rural/urban PHC, clinical IMCI classification, PO uptake, SpO_2_ level measurement (severe and moderate hypoxemia), IMCI+PO classification, IMCI support (on paper/electronic), severe cases (respiratory or non-respiratory) and proportion among severe cases with hospital transfer decided by HCW. Quantitative data were described using means and standard deviations or using median and interquartile ranges and compared using Student’s t- tests. Categorial data were described with proportions with their 95% confidence intervals (CI) and were compared using Pearson χ2 or Fischer exact tests. All analysis were considered statistically significant with p-value less than 0.05.

Then, we compared the main characteristics according to disease severity for the whole sample, and analysed the factors associated with severe disease diagnosis, using a generalized linear mixed regression model with a random country effect. We computed an explanatory model with univariate analysis, then adjusted a full model including the relevant variables or those associated with a p-value <0.20 in univariate analysis. We report adjusted Odds Ratio (aOR) with their 95% CI. Variables explored were: age and sex (forced), mother’s vital status and literacy level, existing income-generating activity, travel time from home to PHC (>30 min), consultation delay (>2 days) since the onset of disease, and type of IMCI support (paper or electronic). A two-tailed p-value of <0.05 was regarded as statistically significant. R software version 4.0.5 was used for all analysis.

### Ethical aspects

The AIRE research protocol, has been approved by each national ethics committee, by the Inserm Institutional Evaluation Ethics Committee (IEEC) and the WHO Ethics Review Committee (WHO-ERC): Comité d’Ethique pour la Recherche en Santé (CERS), Burkina Faso n°2020-4-070; Comité National d’Ethique pour la Recherche en Santé (CNERS), Guinea n°169/CNERS/21; Comité National d’Éthique pour la Santé et les Sciences de la vie (CNESS), Mali n°127/MSDS-CNESS; Comité National d’Ethique pour la Recherche en Santé (CNERS) Niger n°67/2020/CNERS; Inserm IEEC n°20-720; WHO-ERC n° ERC.0003364). This study has been retrospectively registered by the Pan African Clinical Trials Registry on June 15th 2022 under the following Trial registration number: PACTR202206525204526(28).

### Patient and public involvement

This study was conducted using individual data collected with the ethical committee and MOH authorization. Patients were not involved in the design, or conduct, or reporting, or dissemination plans of our research.

## Results

### Flow chart

From June 14^th^, 2021 to June 20^th^ 2022, 39,360 children under-5 attended IMCI consultations in the 16 research PHCs (Flow chart, Figure 1). Among them, 7,760 (19.7%) simple non-respiratory IMCI cases not eligible for PO use were excluded. Among the 31,600 (80.3%) children under-5 eligible for PO use, 15,670 (49.6%) seeking services at night or over the weekends when study data collector where not at the PHC, were not offered the study. Among the 15,930 (50.4%) remaining who were offered the study, 33 (0.2%) families refused, mainly because the child’s accompanying person needed the father’ authorization, and 15,897 (99.7%) were included in the research study with parental consent, of whom 61 (0.4%) were excluded from the analysis for missing IMCI classification or wrong inclusion. Overall, 15,836 (99.6% of those included) were considered in analysis, including 968 (6.1%) neonates (0-59 days) and 14,868 (93.9%) children (2-59 months) (Figure1).

**Figure 1:**
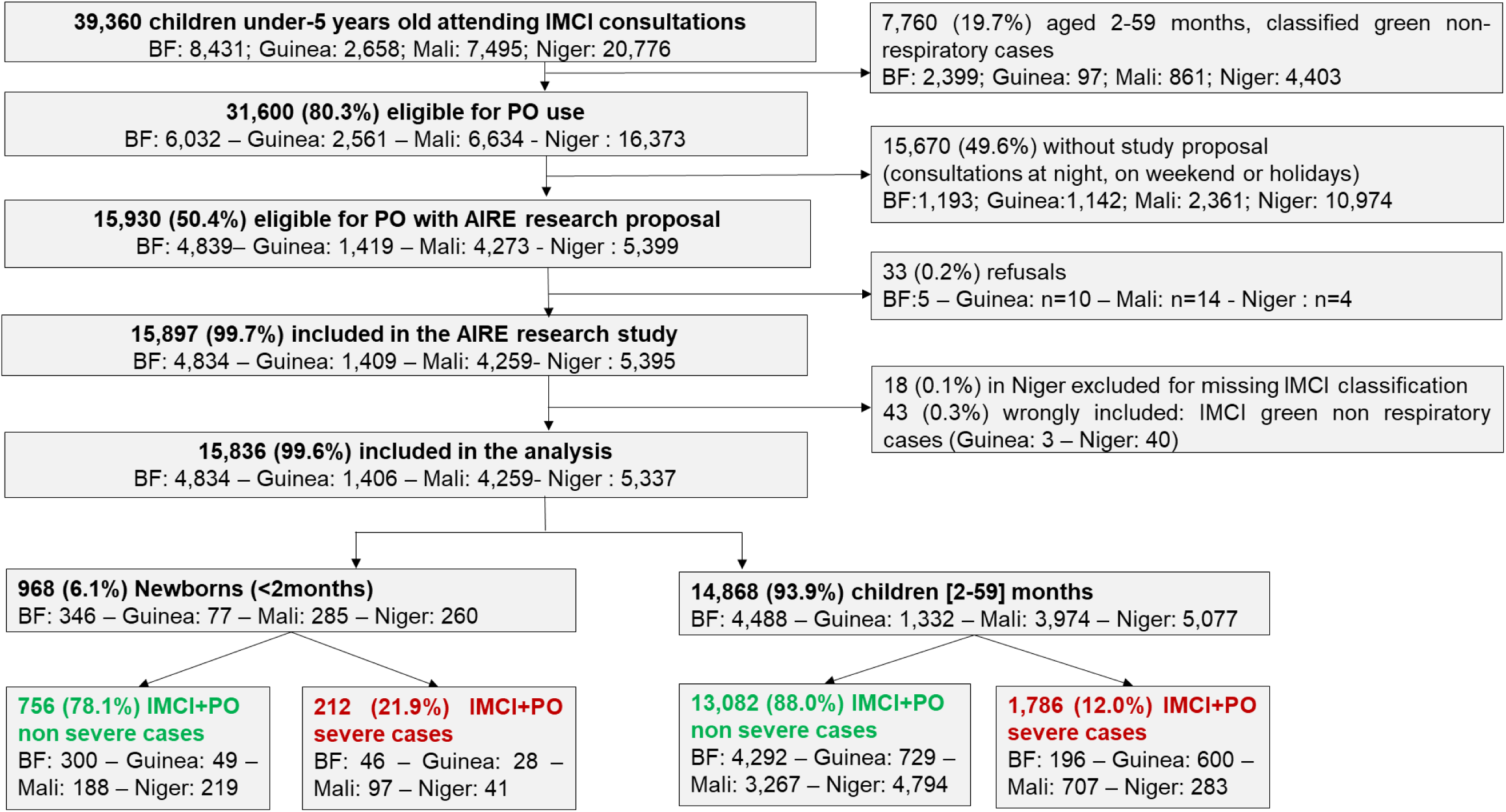
Flowchart of inclusion process in the AIRE research study according to age: June 2021 – June 2022

### Socio-demographics characteristics of outpatients enrolled by country

Table 1 shows the socio-demographic characteristics of the 15,836 children included in the study, globally and by country. Overall, 6.1% of the children included were neonates, ranging from 4.9% in Niger to 7.2% in Burkina Faso. Female sex was under-represented, accounting for 47.2% of the whole sample. The median number of people living under the same roof as the child varied from 5 in Burkina Faso, Guinea and Niger to 9 in Mali. The head of the household had never attended school in 65.1% of cases overall, ranging from 49.1% in Guinea to 82.4% in Burkina Faso. Overall, the child ‘s mother was still alive in 99.6% of cases, and the person accompanying the child on the day of the consultation was either the mother or father in 98.0%, and this person was married or in union in 98.3% of cases. Overall, 81.0% of the children lived less than 30 minutes away from the PHC consulted. In 61.2% of cases, families travel to the PHC by foot, cart, or bike without any expense (ranging from 23.2% in Mali to 83.0% in Burkina Faso), while in 38.6% of cases, families travel by vehicle, motorcycle, or bus (private or public transport) (ranging from 16.0% in Niger to 77.1% in Mali). Children acceded to IMCI consultations in a median delay of 2 days after the onset of symptoms (except in Guinea, where it took 3 days).

**Table 1:**
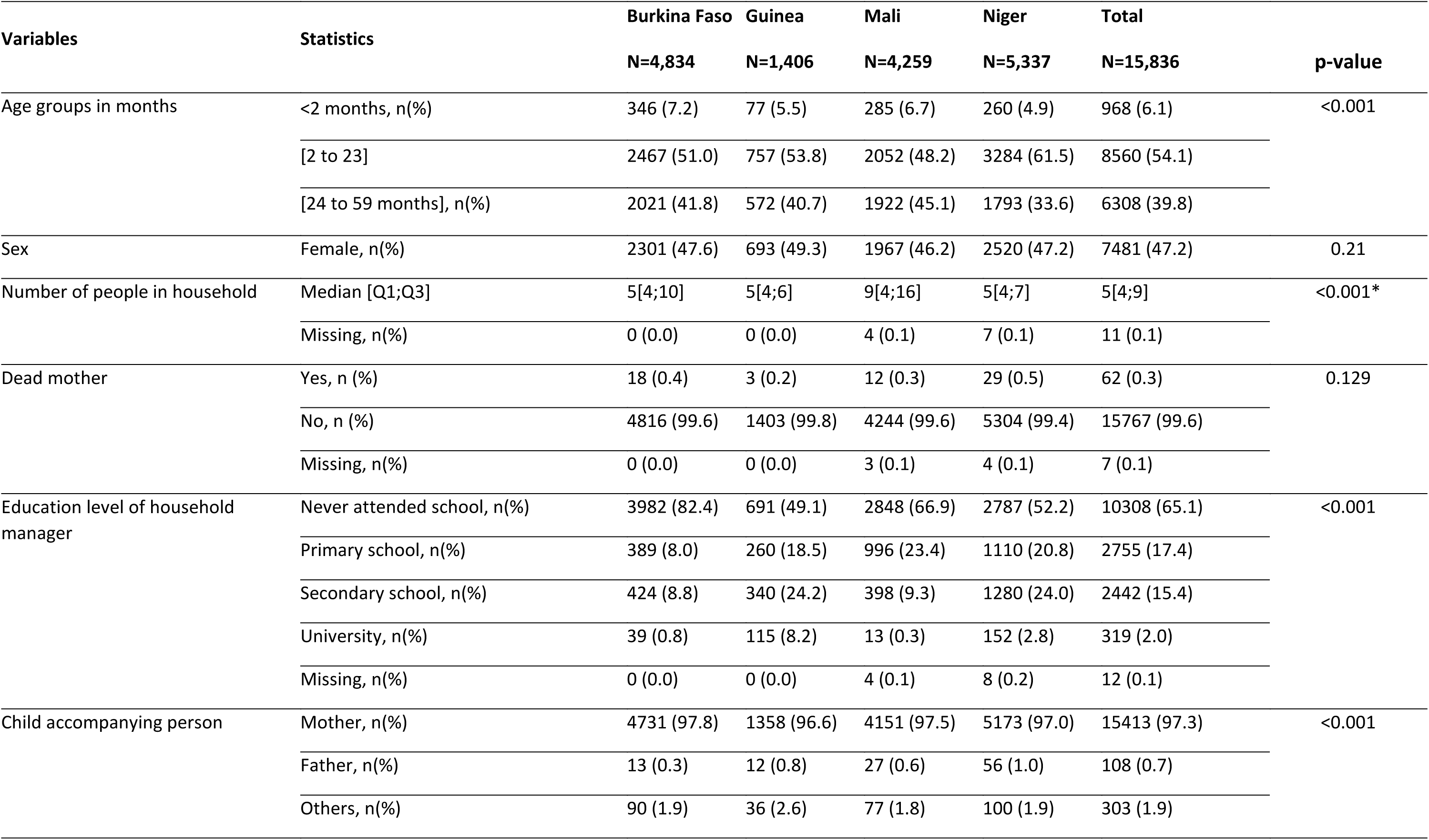
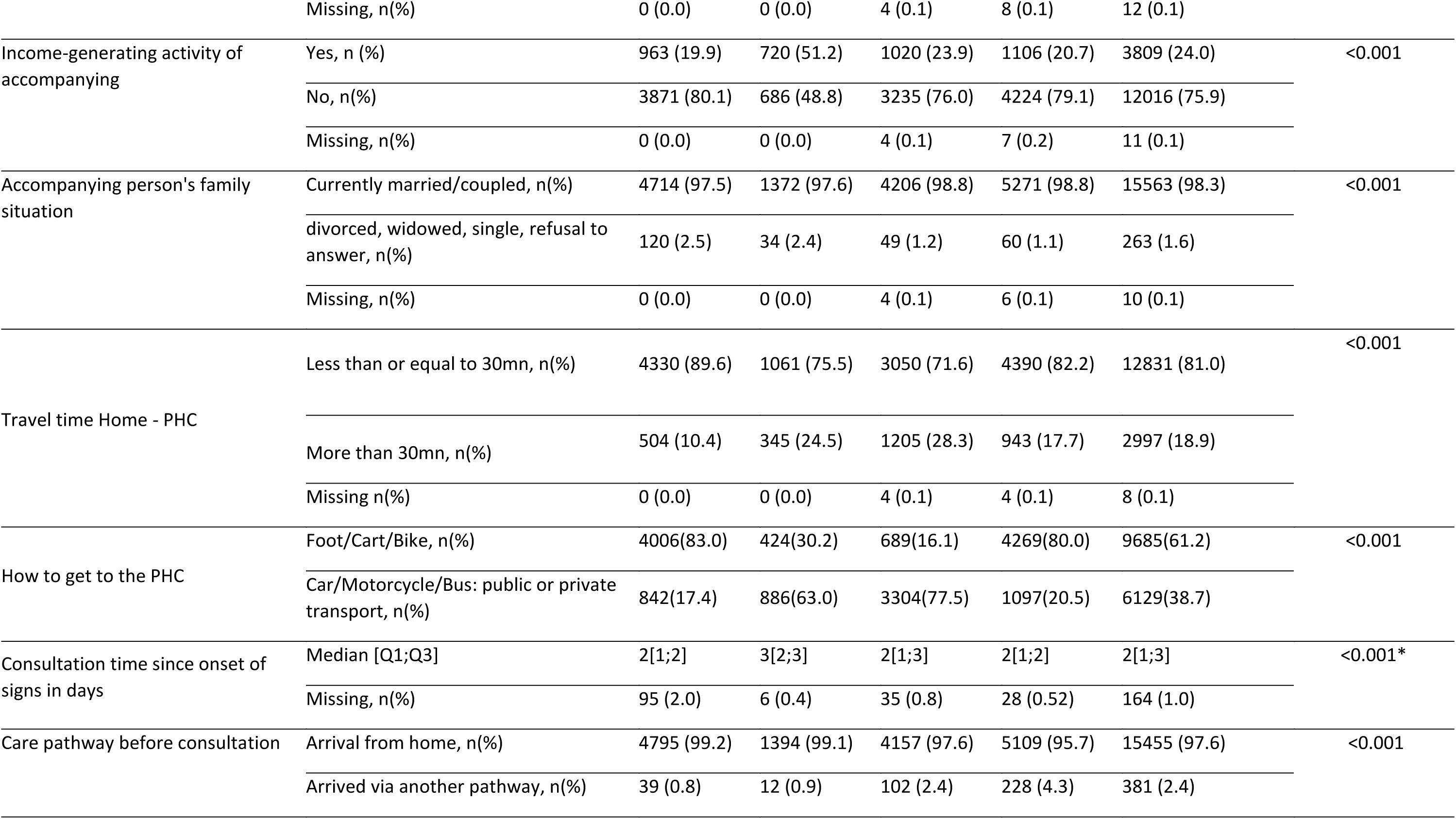
Global and by-country sociodemographic characteristics of IMCI outpatient children enrolled at PHC level in the AIRE research project, June 2021-June 2022, (N=15,836).

| Variables | Statistics | Burkina Faso | Guinea | Mali | Niger | Total | p-value |
| --- | --- | --- | --- | --- | --- | --- | --- |
|  |  | N=4,834 | N=1,406 | N=4,259 | N=5,337 | N=15,836 |  |
| Age groups in months | <2 months, n(%) | 346 (7.2) | 77 (5.5) | 285 (6.7) | 260 (4.9) | 968 (6.1) | <0.001 |
|  | [2 to 23] | 2467 (51.0) | 757 (53.8) | 2052 (48.2) | 3284 (61.5) | 8560 (54.1) |  |
|  | [24 to 59 months], n(%) | 2021 (41.8) | 572 (40.7) | 1922 (45.1) | 1793 (33.6) | 6308 (39.8) |  |
| Sex | Female, n(%) | 2301 (47.6) | 693 (49.3) | 1967 (46.2) | 2520 (47.2) | 7481 (47.2) | 0.21 |
| Number of people in household | Median [Q1;Q3] | 5[4;10] | 5[4;6] | 9[4;16] | 5[4;7] | 5[4;9] | <0.001* |
|  | Missing, n(%) | 0 (0.0) | 0 (0.0) | 4 (0.1) | 7 (0.1) | 11 (0.1) |  |
| Dead mother | Yes, n (%) | 18 (0.4) | 3 (0.2) | 12 (0.3) | 29 (0.5) | 62 (0.3) | 0.129 |
|  | No, n (%) | 4816 (99.6) | 1403 (99.8) | 4244 (99.6) | 5304 (99.4) | 15767 (99.6) |  |
|  | Missing, n(%) | 0 (0.0) | 0 (0.0) | 3 (0.1) | 4 (0.1) | 7 (0.1) |  |
| Education level of household manager | Never attended school, n(%) | 3982 (82.4) | 691 (49.1) | 2848 (66.9) | 2787 (52.2) | 10308 (65.1) | <0.001 |
|  | Primary school, n(%) | 389 (8.0) | 260 (18.5) | 996 (23.4) | 1110 (20.8) | 2755 (17.4) |  |
|  | Secondary school, n(%) | 424 (8.8) | 340 (24.2) | 398 (9.3) | 1280 (24.0) | 2442 (15.4) |  |
|  | University, n(%) | 39 (0.8) | 115 (8.2) | 13 (0.3) | 152 (2.8) | 319 (2.0) |  |
|  | Missing, n(%) | 0 (0.0) | 0 (0.0) | 4 (0.1) | 8 (0.2) | 12 (0.1) |  |
| Child accompanying person | Mother, n(%) | 4731 (97.8) | 1358 (96.6) | 4151 (97.5) | 5173 (97.0) | 15413 (97.3) | <0.001 |
|  | Father, n(%) | 13 (0.3) | 12 (0.8) | 27 (0.6) | 56 (1.0) | 108 (0.7) |  |
|  | Others, n(%) | 90 (1.9) | 36 (2.6) | 77 (1.8) | 100 (1.9) | 303 (1.9) |  |
|  | Missing, n(%) | 0 (0.0) | 0 (0.0) | 4 (0.1) | 8 (0.1) | 12 (0.1) |  |
| Income-generating activity of accompanying | Yes, n (%) | 963 (19.9) | 720 (51.2) | 1020 (23.9) | 1106 (20.7) | 3809 (24.0) | <0.001 |
|  | No, n(%) | 3871 (80.1) | 686 (48.8) | 3235 (76.0) | 4224 (79.1) | 12016 (75.9) |  |
|  | Missing, n(%) | 0 (0.0) | 0 (0.0) | 4 (0.1) | 7 (0.2) | 11 (0.1) |  |
| Accompanying person's family situation | Currently married/coupled, n(%) | 4714 (97.5) | 1372 (97.6) | 4206 (98.8) | 5271 (98.8) | 15563 (98.3) | <0.001 |
|  | divorced, widowed, single, refusal to answer, n(%) | 120 (2.5) | 34 (2.4) | 49 (1.2) | 60 (1.1) | 263 (1.6) |  |
|  | Missing, n(%) | 0 (0.0) | 0 (0.0) | 4 (0.1) | 6 (0.1) | 10 (0.1) |  |
| Travel time Home - PHC | Less than or equal to 30mn, n(%) | 4330 (89.6) | 1061 (75.5) | 3050 (71.6) | 4390 (82.2) | 12831 (81.0) | <0.001 |
|  | More than 30mn, n(%) | 504 (10.4) | 345 (24.5) | 1205 (28.3) | 943 (17.7) | 2997 (18.9) |  |
|  | Missing n(%) | 0 (0.0) | 0 (0.0) | 4 (0.1) | 4 (0.1) | 8 (0.1) |  |
| How to get to the PHC | Foot/Cart/Bike, n(%) | 4006(83.0) | 424(30.2) | 689(16.1) | 4269(80.0) | 9685(61.2) | <0.001 |
|  | Car/Motorcycle/Bus: public or private transport, n(%) | 842(17.4) | 886(63.0) | 3304(77.5) | 1097(20.5) | 6129(38.7) |  |
| Consultation time since onset of signs in days | Median [Q1;Q3] | 2[1;2] | 3[2;3] | 2[1;3] | 2[1;2] | 2[1;3] | <0.001* |
|  | Missing, n(%) | 95 (2.0) | 6 (0.4) | 35 (0.8) | 28 (0.52) | 164 (1.0) |  |
| Care pathway before consultation | Arrival from home, n(%) | 4795 (99.2) | 1394 (99.1) | 4157 (97.6) | 5109 (95.7) | 15455 (97.6) | <0.001 |
|  | Arrived via another pathway, n(%) | 39 (0.8) | 12 (0.9) | 102 (2.4) | 228 (4.3) | 381 (2.4) |  |

### Clinical characteristics, prevalence of severe illness after IMCI+PO classification, and HCW’s decision after IMCI consultations by age and by country

The clinical characteristics of the 15,836 children included and outcomes of the IMCI+PO consultations differed significantly according to age and country (Table 2).

**Table 2:**
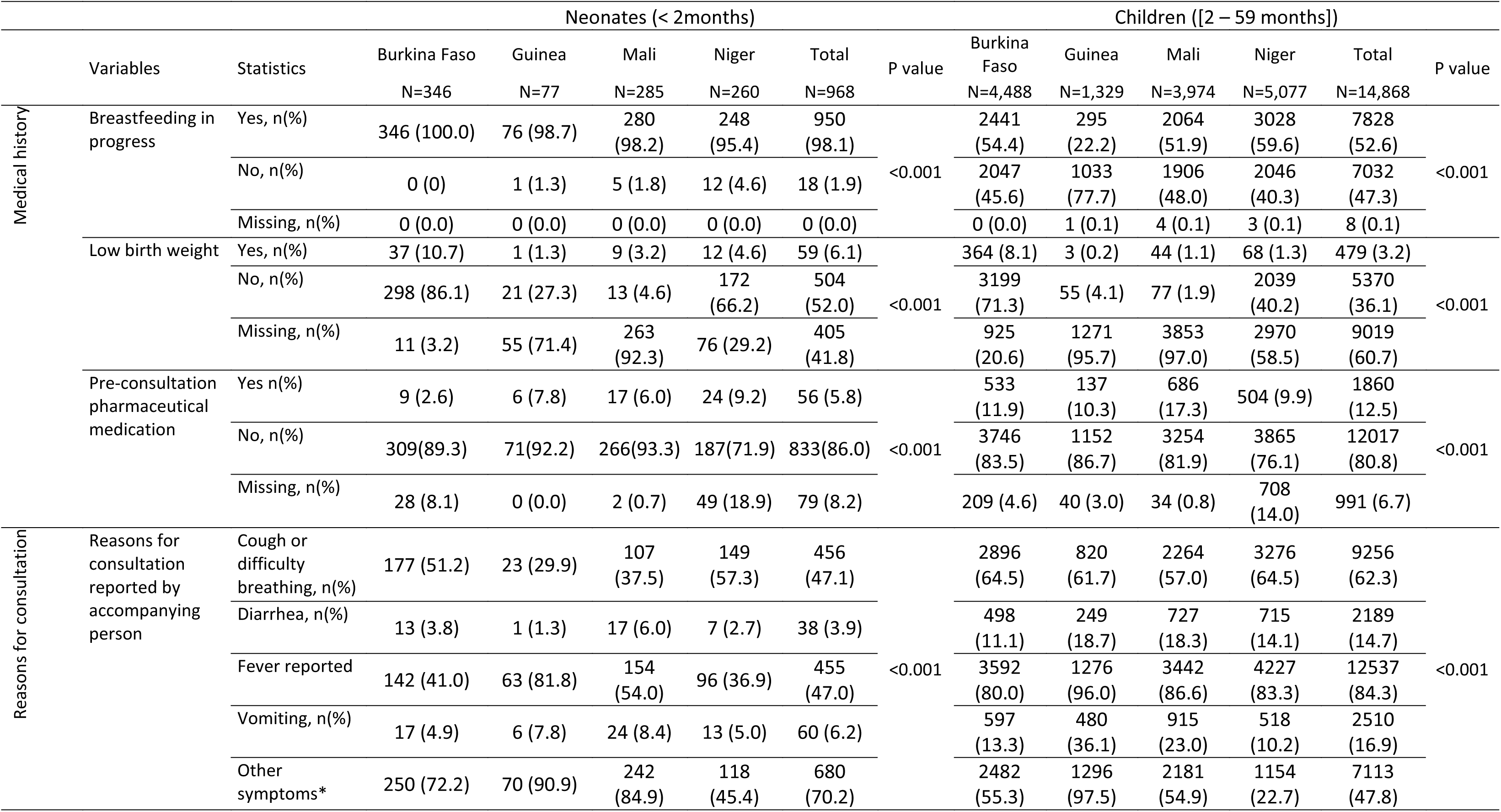
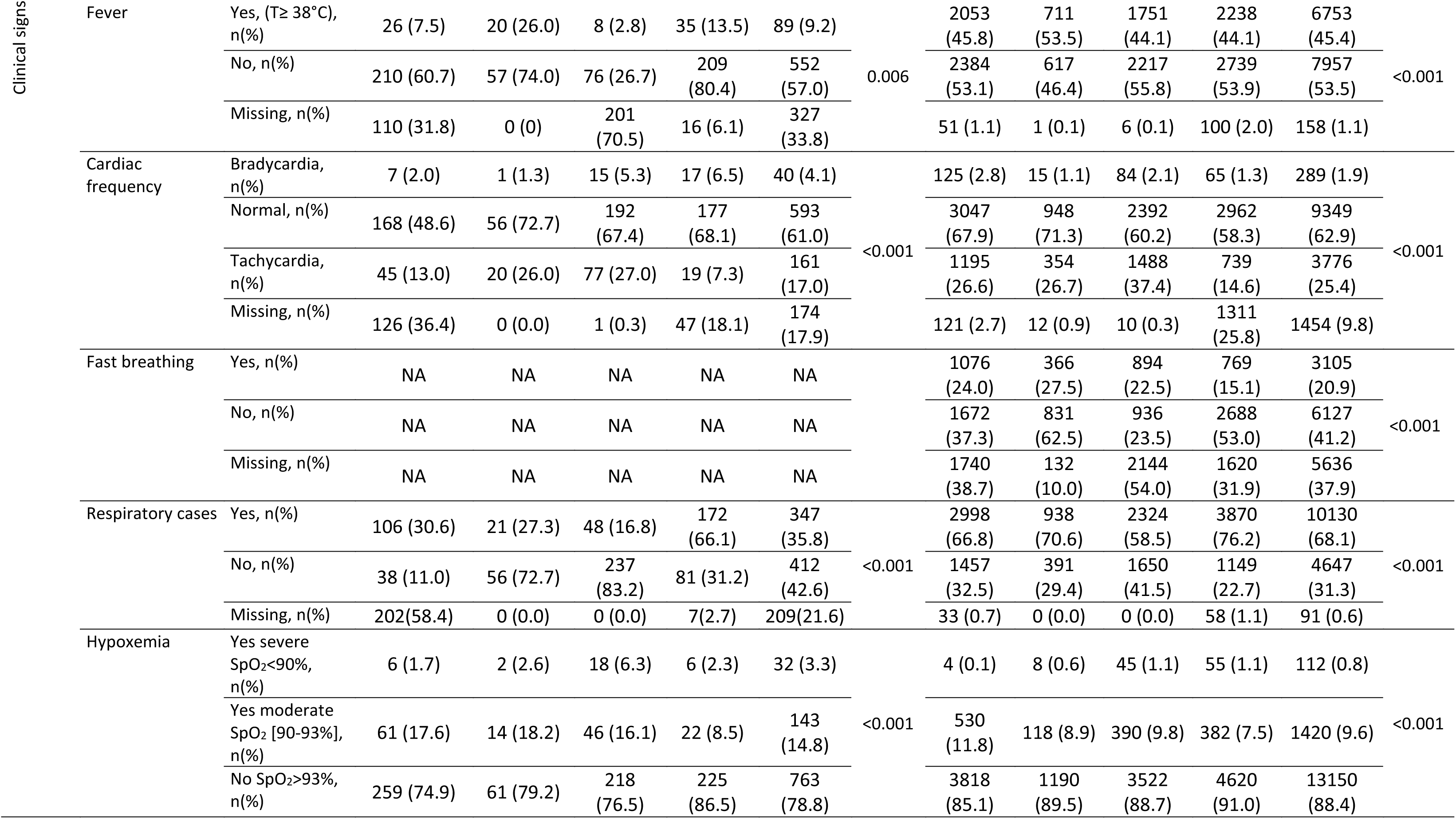
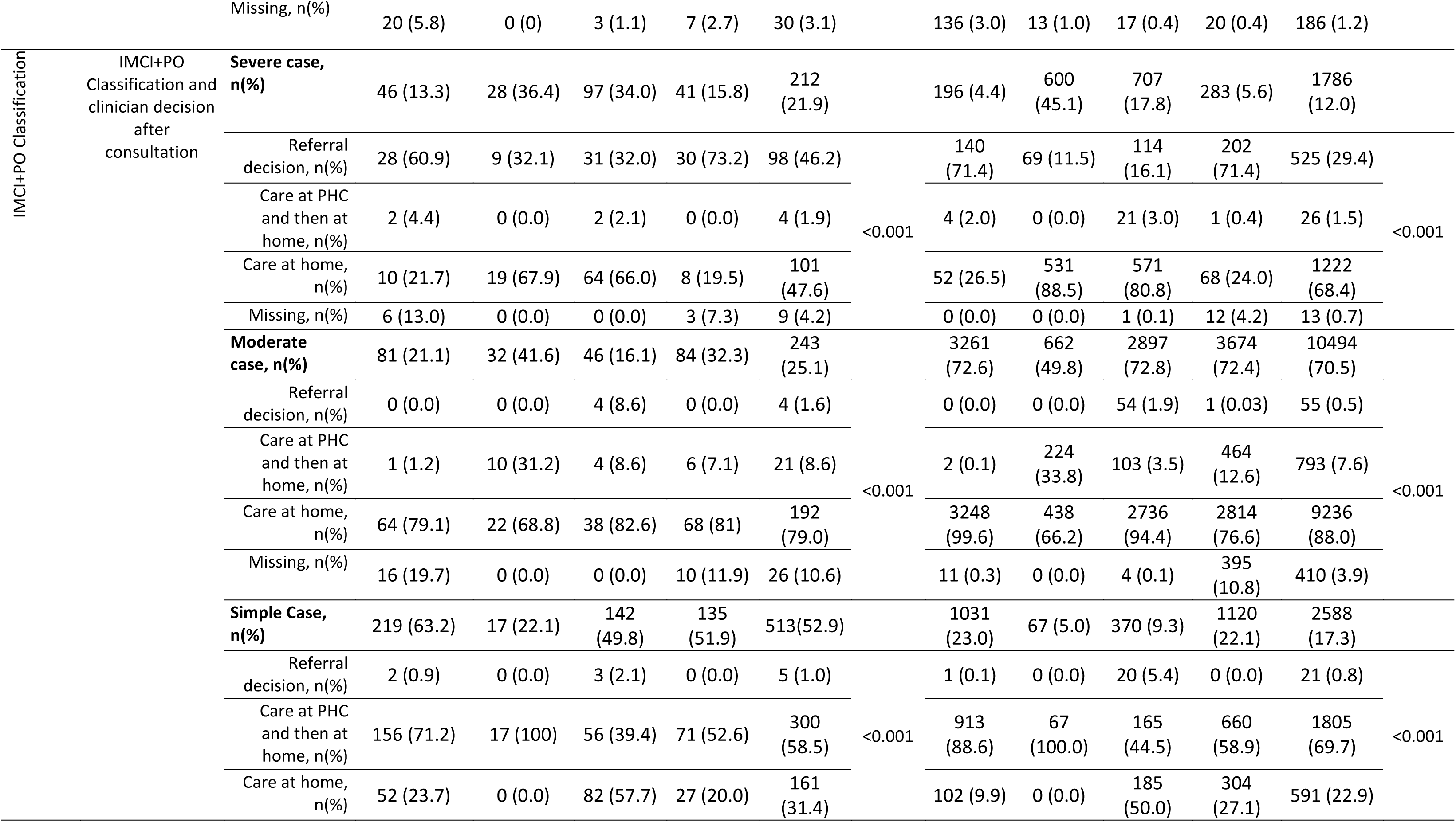
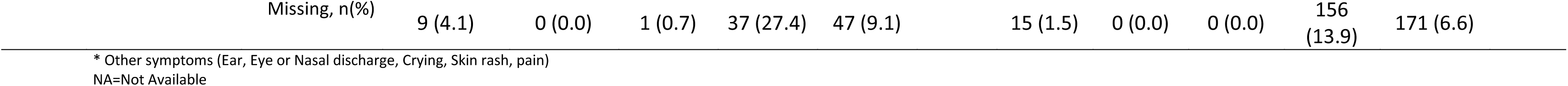
Clinical characteristics of IMCI outpatient neonates (< 2 months) and children (2 to 59 months) enrolled at PHC level in the AIRE project. June 2021-June 2022, (N=15,836)

Among the 968 neonates included, 6.1% had low birth weight (varying significantly from 1.3% in Guinea to 10.7% in Burkina Faso). Breastfeeding in the first months of life was the rule (98.0% were breast-feeding at the time of visit); 5.8% had received medication prior to the IMCI consultation (varying from 2.6% in Burkina Faso to 9.2% in Niger). The main reasons for consultation reported by the accompanying person were cough or respiratory difficulties for 47.1% of all the neonates, reported fever (47.0%), vomiting (6.2%) and diarrhoea (3.9%). After the IMCI consultation, fever (T≥38°C) was measured in 9.2% of the neonates (varying from 2.8% in Mali to 26.0% in Guinea). Bradycardia was noted in 4.1% while fast heart rate (tachycardia) was present in only 17.0%; 35.8% were classified as respiratory cases. Hypoxemia (SpO_2_≤93%) was prevalent in 18.3% of all neonates, 3.3% (95% CI: 2.3 – 4.6) severe hypoxemia and 14.8% (95% CI: 12.6 – 17.2) moderate hypoxemia. Severe hypoxemia varied significantly from 1.7% in Burkina Faso to 6.3% in Mali. After, the IMCI classification using PO, the prevalence of severe cases was estimated at 21.9% (212/968, 95% CI: 19.3 – 24.6), with a substantial variation between countries, 13.3%, 15.8%, 34.0%, and 36.4% in Burkina Faso, Niger, Mali and Guinea, respectively (Table 2). The clinician decision to refer those neonates classified as severe cases was taken overall for 46.2% (95% CI: 39.4 - 53.2), with high between-country heterogeneity: 32.0%, 32.1%, 60.9%, and 73.2% in Mali, Guinea, Burkina Faso and Niger, respectively.

Among the 14,868 children aged 2-59 months included; 12.5% had received medication prior to the IMCI consultation (varying from 9.9% in Niger to 17.3% in Mali). The main reasons for consultation reported by the accompanying person were fever (84.3%), cough or respiratory difficulties (62.3%), followed by vomiting (16.9%) and diarrhoea (14.7%). After the IMCI consultation, clinically diagnosed fever (T≥38°C) was noted in 45.4% of the children (varying from 44.1% in Mali and Niger to 53.5% in Guinea). Sixty eight percent (68.1%) were classified as respiratory cases; hypoxemia was prevalent in 10.4% children; 0.8% (95% CI: 0.6 – 0.9) severe hypoxemia and 9.6% (95% CI: 9.1 – 10) moderate hypoxemia. Severe hypoxemia varied significantly from 0.1% in Burkina Faso to 1.1% in Mali and Niger. Overall using IMCI+PO, 70.5% of children were classified as moderate cases: 49.8% in Guinea and around 72.0%, elsewhere. The prevalence of IMCI+PO severe cases was estimated at 12.0% (95% CI: 11.4 – 12.5), with significantly variations between countries: 4.4%, 5.6%, 17.8% and 45.1% in Burkina Faso, Niger, Mali and Guinea, respectively. At the end of the IMC+PO consultation, HCWs took the decision to refer those classified as severe cases for 29.4% overall (95% CI: 27.3 – 31.6), with high between-country heterogeneity, 11.5% in Guinea, 16.1% in Mali and 71.4% in Burkina Faso and Niger (Table 2).

### Factors associated with the identification of severe illness using IMCI+PO

Overall, among the 15,836 children included, 1,998 (12.6%) were classified as severe cases using IMCI+PO classification, including 212 (10.6%) neonates, and 1,786 (89.4%) older children (Figure 1). Prevalence of severe cases varied significantly between countries, estimated at 5.0% (95% CI: 4.4 - 5.7) in Burkina Faso, 6.1% (95% CI: 5.4 - 6.7) in Niger, 18.9% (95% CI: 17.7 - 20.1) in Mali, and 44.7% (95% CI: 42.0 - 47.3) in Guinea. The distribution of IMCI+PO classification by country is shown in Figure 2.

**Figure2:**
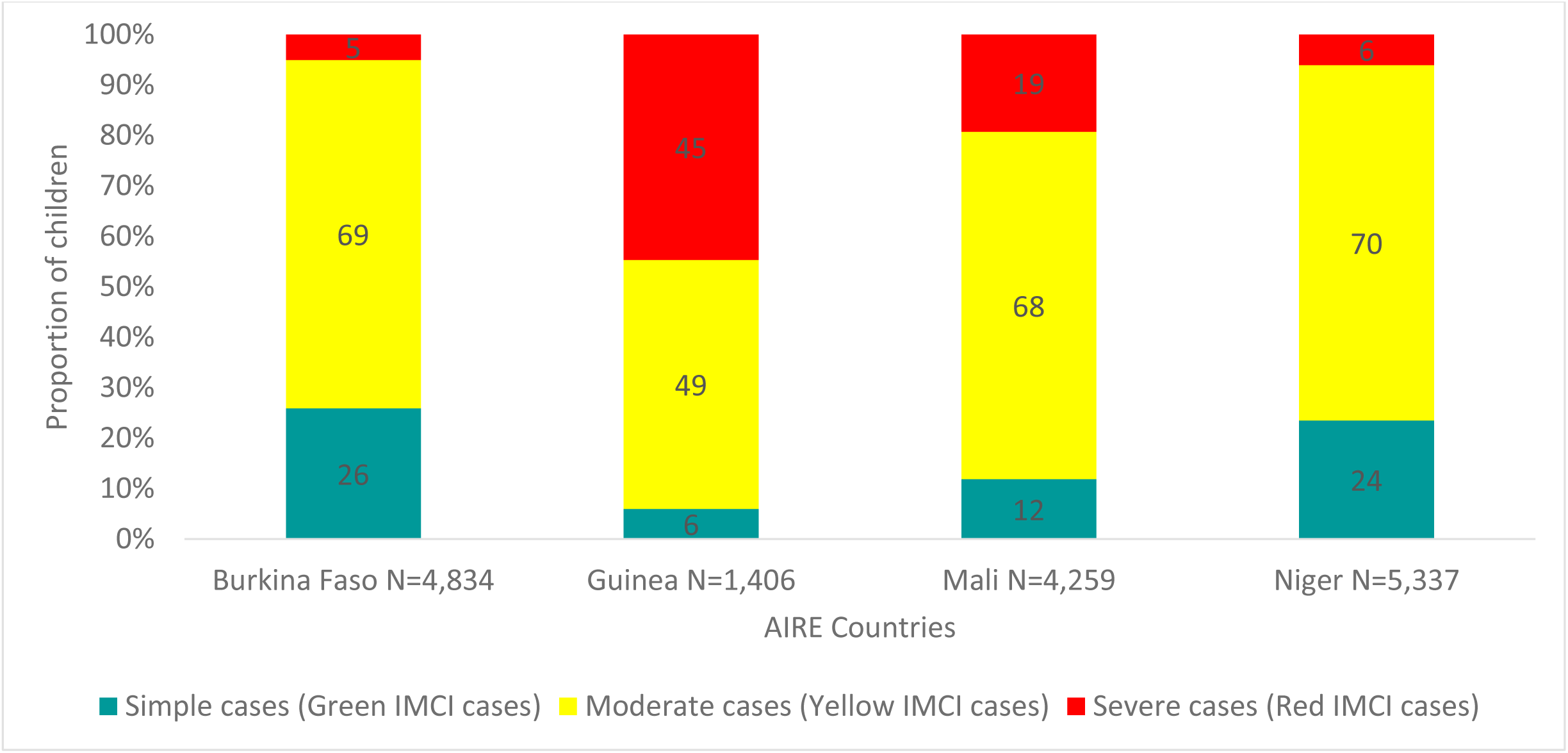
IMCI Classification using PO of children under-5 years of age included at PHC level in the four AIRE countries project, June 2021 – June 2022

Children aged <2 months and 24-59 months were more likely to be diagnosed as severe cases, 10.6% and 47.0%, respectively than those aged between 2 and 23 months (42.3%) (p <0.001) (table 3). Children whose household responsible was illiterate, had no income generating activity, living at more than 30 min travel from the PHC, presenting with signs more than 2 days since the onset of symptoms or through another pathway of care were significantly more likely to be diagnosed as severe cases. Severe cases compared to non-severe cases were significantly more likely to present with fever measured (p <0.001), severe hypoxemia (p <0.001), and non-respiratory symptoms (p <0.001).

**Table 3:**
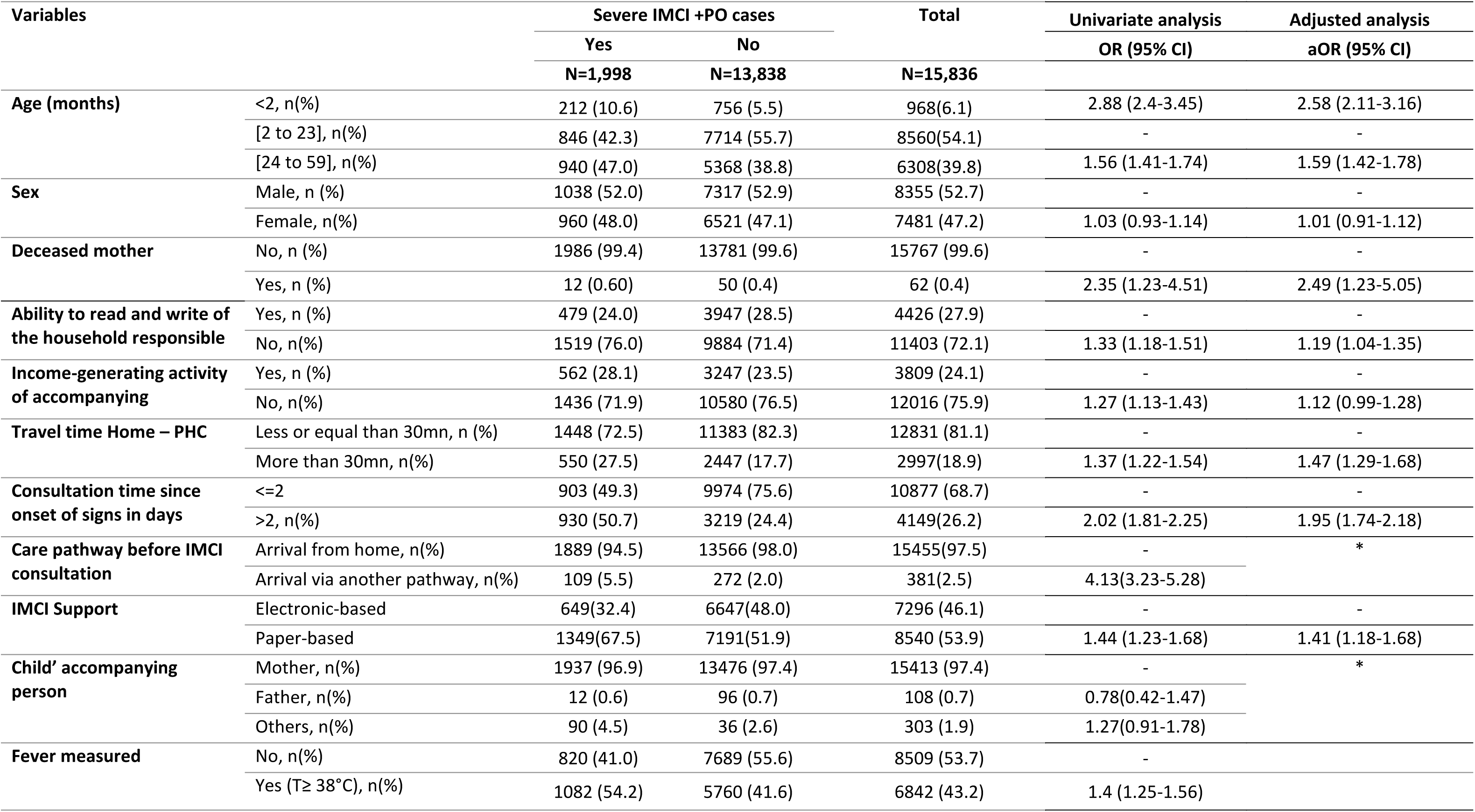
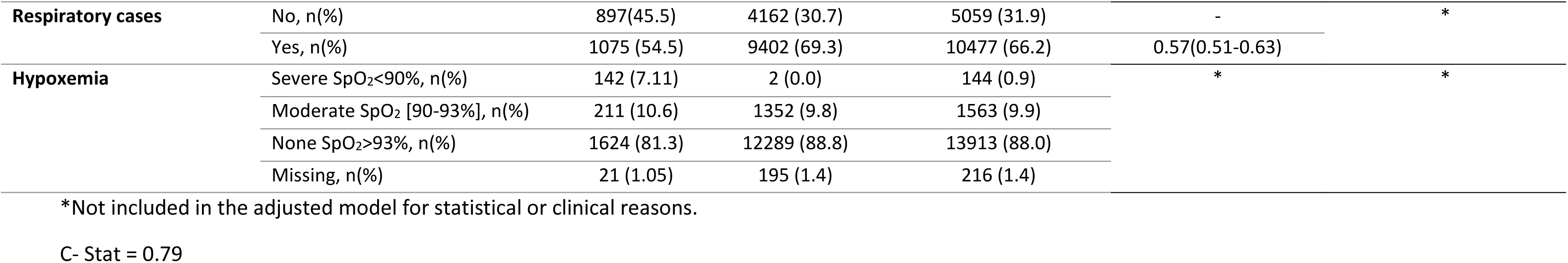
Sociodemographic and clinical distribution and factors associated to illness severity of IMCI children enrolled at PHC in the AIRE project, using a logistic regression Model with a random country effect, AIRE project June 2021 – June 2022 (1,998 vs 13,838)

| Variables |  | Severe IMCI +PO cases |  | Total<br>N=15,836 | Univariate analysis | Adjusted analysis |
| --- | --- | --- | --- | --- | --- | --- |
|  |  | Yes | No |  | OR (95% CI) | aOR (95% CI) |
|  |  | N=1,998 | N=13,838 |  |  |  |
| Age (months) | <2, n(%) | 212 (10.6) | 756 (5.5) | 968(6.1) | 2.88 (2.4-3.45) | 2.58 (2.11-3.16) |
|  | [2 to 23], n(%) | 846 (42.3) | 7714 (55.7) | 8560(54.1) | - | - |
|  | [24 to 59], n(%) | 940 (47.0) | 5368 (38.8) | 6308(39.8) | 1.56 (1.41-1.74) | 1.59 (1.42-1.78) |
| Sex | Male, n (%) | 1038 (52.0) | 7317 (52.9) | 8355 (52.7) | - | - |
|  | Female, n(%) | 960 (48.0) | 6521 (47.1) | 7481 (47.2) | 1.03 (0.93-1.14) | 1.01 (0.91-1.12) |
| Deceased mother | No, n (%) | 1986 (99.4) | 13781 (99.6) | 15767 (99.6) | - | - |
|  | Yes, n (%) | 12 (0.60) | 50 (0.4) | 62 (0.4) | 2.35 (1.23-4.51) | 2.49 (1.23-5.05) |
| Ability to read and write of the household responsible | Yes, n (%) | 479 (24.0) | 3947 (28.5) | 4426 (27.9) | - | - |
|  | No, n(%) | 1519 (76.0) | 9884 (71.4) | 11403 (72.1) | 1.33 (1.18-1.51) | 1.19 (1.04-1.35) |
| Income-generating activity of accompanying | Yes, n (%) | 562 (28.1) | 3247 (23.5) | 3809 (24.1) | - | - |
|  | No, n(%) | 1436 (71.9) | 10580 (76.5) | 12016 (75.9) | 1.27 (1.13-1.43) | 1.12 (0.99-1.28) |
| Travel time Home – PHC | Less or equal than 30mn, n (%) | 1448 (72.5) | 11383 (82.3) | 12831 (81.1) | - | - |
|  | More than 30mn, n(%) | 550 (27.5) | 2447 (17.7) | 2997(18.9) | 1.37 (1.22-1.54) | 1.47 (1.29-1.68) |
| Consultation time since onset of signs in days | <=2 | 903 (49.3) | 9974 (75.6) | 10877 (68.7) | - | - |
|  | >2, n(%) | 930 (50.7) | 3219 (24.4) | 4149(26.2) | 2.02 (1.81-2.25) | 1.95 (1.74-2.18) |
| Care pathway before IMCI consultation | Arrival from home, n(%) | 1889 (94.5) | 13566 (98.0) | 15455(97.5) | - | * |
|  | Arrival via another pathway, n(%) | 109 (5.5) | 272 (2.0) | 381(2.5) | 4.13(3.23-5.28) |  |
| IMCI Support | Electronic-based | 649(32.4) | 6647(48.0) | 7296 (46.1) | - | - |
|  | Paper-based | 1349(67.5) | 7191(51.9) | 8540 (53.9) | 1.44 (1.23-1.68) | 1.41 (1.18-1.68) |
| Child' accompanying person | Mother, n(%) | 1937 (96.9) | 13476 (97.4) | 15413 (97.4) | - | * |
|  | Father, n(%) | 12 (0.6) | 96 (0.7) | 108 (0.7) | 0.78(0.42-1.47) |  |
|  | Others, n(%) | 90 (4.5) | 36 (2.6) | 303 (1.9) | 1.27(0.91-1.78) |  |
| Fever measured | No, n(%) | 820 (41.0) | 7689 (55.6) | 8509 (53.7) | - |  |
|  | Yes (T≥ 38°C), n(%) | 1082 (54.2) | 5760 (41.6) | 6842 (43.2) | 1.4 (1.25-1.56) |  |
| **Respiratory cases** | No, n(%) | 897(45.5) | 4162 (30.7) | 5059 (31.9) | - | * |
|  | Yes, n(%) | 1075 (54.5) | 9402 (69.3) | 10477 (66.2) | 0.57(0.51-0.63) |  |
| **Hypoxemia** | Severe SpO<sub>2</sub><90%, n(%) | 142 (7.11) | 2 (0.0) | 144 (0.9) | * | * |
|  | Moderate SpO<sub>2</sub> [90-93%], n(%) | 211 (10.6) | 1352 (9.8) | 1563 (9.9) |  |  |
|  | None SpO<sub>2</sub>>93%, n(%) | 1624 (81.3) | 12289 (88.8) | 13913 (88.0) |  |  |
|  | Missing, n(%) | 21 (1.05) | 195 (1.4) | 216 (1.4) |  |  |

\*Not included in the adjusted model for statistical or clinical reasons.
C- Stat = 0.79

When exploring social health factors associated with disease severity in a global generalised mixed linear model with a random country effect adjusted on sex, income generating activity of household responsible, the following factors were independently associated with an increased risk of illness severity (Table3): children aged <2 months (aOR: 2.58: 95% CI: 2.11-3.16), and children aged from 24 to 59 months (aOR:1.59, 95% CI: 1.42-1.78) compared to those aged 2-23 months, those whose mother is deceased (aOR:2.49; 95% CI: 1.23-5.05), the illiteracy of the child’s accompanying person (aOR=1.12; 95% CI: 1.04-1.35), consultation delay more than 2 days since the onset of symptoms (aOR=1.95; 95% CI: 1.74-2.18), travel time from home to PHC >30 minutes (aOR=1.47; 95% CI: 1.29-1.68), and using a paper-based IMCI vs. electronic-IMCI (aOR=1.41; 95% CI: 1.18-1.68).

We also carried out the same analyses for each country (supplementary tables 1-4), and factors associated with the identification of severe cases varied from country to country.

In Burkina Faso, adjusted risk factors independently associated with severe illness were: age <2 months (aOR=4.94; 95% CI: 3.13-7.69) and between 2 and 5 years old (aOR=2.18; 95% CI: 1.56-3.07) and having a consultation delay of more than 2 days (aOR=2.17; 95% CI: 1.54-3.02).

In Mali, these factors were age under-2 months (aOR=3.18; 95% CI: 2.35-4.27) and children aged 2 to 5 years (aOR=1.83; 95% CI: 1.54-2.18), consultation time of more than 2 days (aOR=1.36; 95% CI: 1.15- 1.61), travel time from home to the PHC >30 minutes (aOR=1.25; 95% CI: 1.02-1.52), and the use of paper-based IMCI compared to electronic IMCI (on tablet) (aOR=1.40; 95% CI: 1.16-1.69).

In Niger, the factors were the age of the child less than 2 months (aOR=1.97; 95% CI: 1.25-3.02), the illiteracy of the child’s accompanying person (aOR=2.43; 95% CI: 1.81-3.30), the consultation time of more than 2 days (aOR=4.44; 95% CI: 3.46-5.71) and a travel time from home to PHC of more than 30 minutes (aOR=1.62; 95% CI: 1.23-2.11).

Finally, in Guinea, an age of 2 to 5 years (aOR=1.76; 95% CI: 1.40-2.20), a consultation delay of more than 2 days (aOR=1.71; 95% CI: 1.37-2.14), and a travel time of more than 30 minutes from home to the PHC (aOR=1.66; 95% CI: 1.29-2.14) were the independent risk factor associated to the disease severity. In Guinea, the illiteracy of the child’s accompanying person was found negatively associated with the identification of severe cases (aOR=0.75; 95% CI: 0.60-0.94).

## Discussion

Our study provides original epidemiological indicators of severe illnesses using IMCI+PO classification among outpatient children attending IMCI consultations at frontline for four West African countries. These data were collected from children under-5 years of age, including neonates, and using standardized tools allowing for between countries and age groups comparisons. Some important findings have been found. First, the prevalence of children classified as severe cases using IMCI+PO was overall high, but significantly heterogeneous between countries, varying from 5.0% in Burkina Faso to 44.6% in Guinea. Second, overall prevalence of severe cases was significantly twice higher in neonates (21.9%) than in children aged 2-59 months (12.0%). Similarly, the prevalence of severe hypoxemia was higher in neonates (3.3%) than in older children (0.8%). Third, among the social health factors globally associated with severe disease at PHC, young age (<2 months), older age (2 to 5 years), maternal death, or illiteracy were identified, as well as variables indicating a delayed access to care (delay> 2 days after the onset of symptoms, and travel duration to get to the PHC >30 min travel time from home to PHC). Fourth, in Mali where comparison of IMCI support was feasible, using a paper-based IMCI seems also to independently increase of +40% the risk of being classified as severe compared to e-IMCI. Finally, the clinician decision at PHC to refer to hospital children classified as severe cases using IMCI+PO was overall insufficient, but also heterogenous according to countries and ages, higher for neonates (46.2%, from 32.0% in Mali to 73.2% in Niger) than for older children (29.4%, from 11.5% in Guinea to 71.5% in Burkina Faso).

A few studies have reported the PHC-based prevalence of severe cases using IMCI+PO among children under-5 years of age in LMICs, and to our knowledge, none had been done in West Africa. A study carried out in Papua New Guinea in 2019 reported a prevalence of severe IMCI+PO cases of 8.3% among the 1663 outpatient children aged 3-27 months seen in PHCs (32). Elsewhere, it was mainly the prevalence of severe pneumonia that was reported using IMCI+PO: McCollum and colleagues, for instance reported a community-based prevalence of severe pneumonia at 8.1% in Bangladesh in 2023 (33); in Ethiopia the prevalence of severe pneumonia was at 15.9% (34). Our estimates of prevalence of severe cases lie within these previous estimates.

The wide disparity observed in the prevalence of severe cases between the four countries may be explained by several structural and individual factors. First, access to care is impacted by the out-of-pocket payments policy in place in each country, as we have demonstrated elsewhere (35). Indeed, Burkina Faso and Niger apply a total free care policy for all children under-5 years of age (36,37): we assume that this policy has partly facilitated an earlier access to care for sick children explaining the lowest prevalence of severe cases in these countries. Mali and Guinea are countries that apply a policy of partial free healthcare for children under-5 years of age (38–40), targeting only few diseases (malaria, malnutrition, HIV and tuberculosis) that have contributed to highest observed prevalence of severe cases among sick children attending IMCI consultations. Second, geographical difficulty in accessing PHCs for population, such as very rugged terrain or waterways in Guinea, or long distances in Mali, are real obstacles in accessing care. This may explain the median 2-day delay between the onset of signs and attendance of PHC, reaching 3 days in Guinea, while the proximity of PHCs to the population in Niamey may explain the lower prevalence of severe cases in Niger (29). Third, the IMCI classification disease management protocols could have played a role. Although the 2014 IMCI guidelines proposed by the WHO (11,12) have been adopted by all countries, there are nevertheless differences between countries that could explain the high prevalence of severe cases observed in Guinea. There the “Chest indrawing” sign is of severity, whereas in Mali, Burkina Faso and Niger, this sign classifies children as moderate cases. Finally, we found that the use of paper-based IMCI was an independent factor associated with a greater (+40%) identification of severe cases compared to electronic IMCI. In fact, the electronic tool favours a systematic and holistic assessment of child health, leading to a more accurate diagnosis, as reported elsewhere (41). This can explain that the use of the paper format, given the number of documents to be filled in manually, may increase the clinician risk of error in assessing child health, in the way of overestimating severity.

In our study, the overall prevalence of severe hypoxemia (SpO_2_< 90%) in all sick children (except simple non-respiratory cases) attending IMCI consultations at AIRE PHCs, was estimated around 0.9% (95% CI: 0.8 - 1.1). This estimate is compatible with other studies conducted in outpatient clinics, in Papua New-Guinea in 2019 (32) or in Uganda in 2022 (25). Consistently, it was lower than other studies that have estimated its prevalence in population subgroups such as severe pneumonia, severe malaria or specific conditions of severe childhood illness: 5.7% (in 2013) then 8.3% (in 2017) in Malawi (42,43); 27.9% in Mozambique (44); or according to several systematic reviews they varied between 5.9% to 62.5% (45), Subhi *et al.* estimated at 13.3% in 2009 (21) or 31% with Rahman *et al.* (46). In our study, the prevalence of severe hypoxemia was significantly higher in neonates than in older children. This finding was also consistent with other studies reported: McCollum *et al.* has also observed this in Malawi in 2017, with a prevalence of severe hypoxemia estimated at 11.4% for children under-5 months, 8.4% for 6-23 months and 4.7% for 24-59 months (43); as did Graham *et al.* in Nigeria in 2019, with 22.2% for neonates and 10.2% for children under-15 (23). The same observation was made in the literature review made by Subhi *et al.* with the conclusion that one in five neonates was hypoxemic (21). This higher prevalence compared to older children could be explained by the immaturity or physiological fragility of neonates.

The global adjusted modelling analysis of social health factors associated with disease severity highlights several key factors allowing to specifically target at risk populations that would be eligible for focused community interventions: neonates or children aged between 2 and 5 years, those whose mother have died, illiteracy of the child’s accompanying person, and those with accessibility barriers to care (consultation time of more than 2 days, travel time from home to PHC of more than 30 minutes). These factors identified confirm previous evidence, particularly the fragility of neonates (23), the influence of maternal health on child health (47) especially maternal death, and the problem of the delay with high distances between homes and PHCs (48–54).

Of note, the decision to transfer severe cases identified at PHC to the district hospital as recommended by the IMCI guidelines was not taken systematically by clinicians. They took the decision to refer severe cases preferably for neonates (46.2%) than for children aged 2-59 months (29.4%). Several factors may explain this gap. It can be related to the lack of training of HCW in the PHC. As observed in the AIRE PHCs description, there were mainly nurses (81%) and doctors (17%), and the IMCI training/refresher courses were not regular (29). As a result, their level of qualification and skills was generally low and despite the implementation of standardized guidelines such as IMCI guidelines or protocols for the management of specific pathologies, children are not properly well-managed (20). In this context, HCW may not always trust the severity outcome proposed by the algorithm when using IMCI symptom-based algorithms without other available and accurate diagnostic tests (except for mRDT). This would favour the decision not to refer these severe cases. Nevertheless, the proportion of severe cases with referral decision, and their outcome although low, has been improved by the use of the PO in our study, as reported elsewhere (55), and consistently to other studies (30,55–57). Finally, the parents fear to be exposed to unaffordable expenses could also have influenced the HCW’s decision, who did not opt for hospital referral. It can also be linked to major geographical barriers, such as the long distances involved, and to the issue of security, particularly in Burkina Faso, Mali and Niger.

Our study has several limitations. First, our definition of severe cases was based on the criteria of severity according to the IMCI+PO guidelines. These are symptom-based, complex and imperfect guidelines, that are largely operator-dependent, and can therefore lead to inaccurate diagnoses of severe cases, since consultations are carried out by the established HCW at PHC level, without etiological diagnostic tools other than mRDT. In addition, the adaptation of the version proposed by the WHO in each country presents slight differences, particularly in Guinea, and more specifically in the respiratory disease block. These differences in IMCI guidelines application are key factors to be considered when comparing the results between the different countries. Nevertheless, the standardised inclusion criteria and classification procedures used by IMCI provides reliable estimates of severe cases and their correlates within each country. Furthermore, another limitation concerns the representativeness of the severe cases included in our study. Indeed, during the data collection period, our research teams were only present on site on working days, and not at night, at weekends or on public holidays, and sometime for security reasons mainly in Mali and Burkina Faso. This selection bias may have conducted to underestimate the prevalence of severe illnesses, assuming the fact that night and weekend attendance could concentrate more severe cases. To better investigate this potential bias, we have conducted an ancillary study describing the care pathway through PHC level or other itineraries of critically ill children aged 0-5 years arriving at the AIRE district hospitals in the four intervention countries(58).

One of the strengths of our study is that the data collected was standardised between countries, of good quality, with very few missing data overall, which made it possible to compare results between countries. We thus provide original estimates of ambulatory care at the peripheral level of the health pyramid, whereas most of the data in the literature comes from hospitalised children. Our data will be useful for assessing the quality of care for sick children at the primary healthcare level in these West African countries, and for guiding national policies. They will also be useful for estimating the added value of integrating PO into IMCI guidelines in this context, which will be the subject of a specific study(55).

## Conclusion

Our study highlights the high prevalence of severe cases identified at PHC level through the routine use of the PO integrated into IMCI at PHC level in four West African countries, and more specifically among neonates. Prevalence of severe cases were heterogeneous between countries. Although, the structural and social health factors explaining heterogeneity in severe illness prevalence were not all captured, we highlighted the vulnerability of neonates, and children exposed to their mother death, household illiteracy and those with a lack of accessibility to PHC. Improving an earlier access to care remains needed in West Africa to save lives. Finally, decisions to transfer to hospital for better care were made for only 3 out of 10 children identified with a severe illness, so improving the care management of severe cases at frontline remains also a challenge for West African healthcare systems to meet the Sustainable Development Goal 3.

## Data Availability

All data produced in the present study are available upon reasonable request to the authors

## Supplementary materials

**Supplementary table 1:** Factors associated to severity of cases identified at PHC in Burkina Faso using a logistic regression model, AIRE research, June 2021 – June 2022 (n= 242 Vs 4,592)

| Variables | levels | No | Yes | Univariate analysis | Adjusted analysis |
| --- | --- | --- | --- | --- | --- |
|  |  |  |  | OR (95% CI, p value) | aOR (95% CI, p value) |
| Age | 2-23 months | 2396 (52.2) | 71 (29.3) | - | - |
|  | <2 months | 300 (6.5) | 46 (19.0) | 5.17 (3.49-7.62, p<0.001) | 4.94 (3.13-7.69, p<0.001) |
|  | 24-59 months | 1896 (41.3) | 125 (51.7) | 2.22 (1.66-3.01, p<0.001) | 2.18 (1.56-3.07, p<0.001) |
| Sex | Male | 2396 (52.2) | 137 (56.6) | - | - |
|  | Female | 2196 (47.8) | 105 (43.4) | 0.84 (0.64-1.08, p=0.179) | 0.76 (0.56-1.03, p=0.076) |
| Mother deceased | No | 4576 (99.7) | 240 (99.2) | - | - |
|  | Yes | 16 (0.3) | 2 (0.8) | 2.38 (0.38-8.44, p=0.249) | - |
| Can read and write | Yes | 726 (15.8) | 27 (11.2) | - | - |
|  | No | 3866 (84.2) | 215 (88.8) | 1.50 (1.01-2.30, p=0.053) | 1.29 (0.83-2.11, p=0.280) |
| Income-generating activity | Yes | 923 (20.1) | 40 (16.5) | - | - |
|  | No | 3669 (79.9) | 202 (83.5) | 1.27 (0.91-1.82, p=0.176) | 1.17 (0.79-1.79, p=0.447) |
| Consultation time in days | <=2 | 3730 (84.8) | 132 (71.7) | - | - |
|  | >2 | 666 (15.2) | 52 (28.3) | 2.21 (1.57-3.05, p<0.001) | 2.17 (1.54-3.02, p<0.001) |
| Travel time from home to PHC | <=30 mn | 4114 (89.6) | 216 (89.3) | - | - |
|  | >30mn | 478 (10.4) | 26 (10.7) | 1.04 (0.67-1.54, p=0.868) | 1.23 (0.75-1.91, p=0.382) |

C- Stat = 0.67

**Supplementary table 2:**
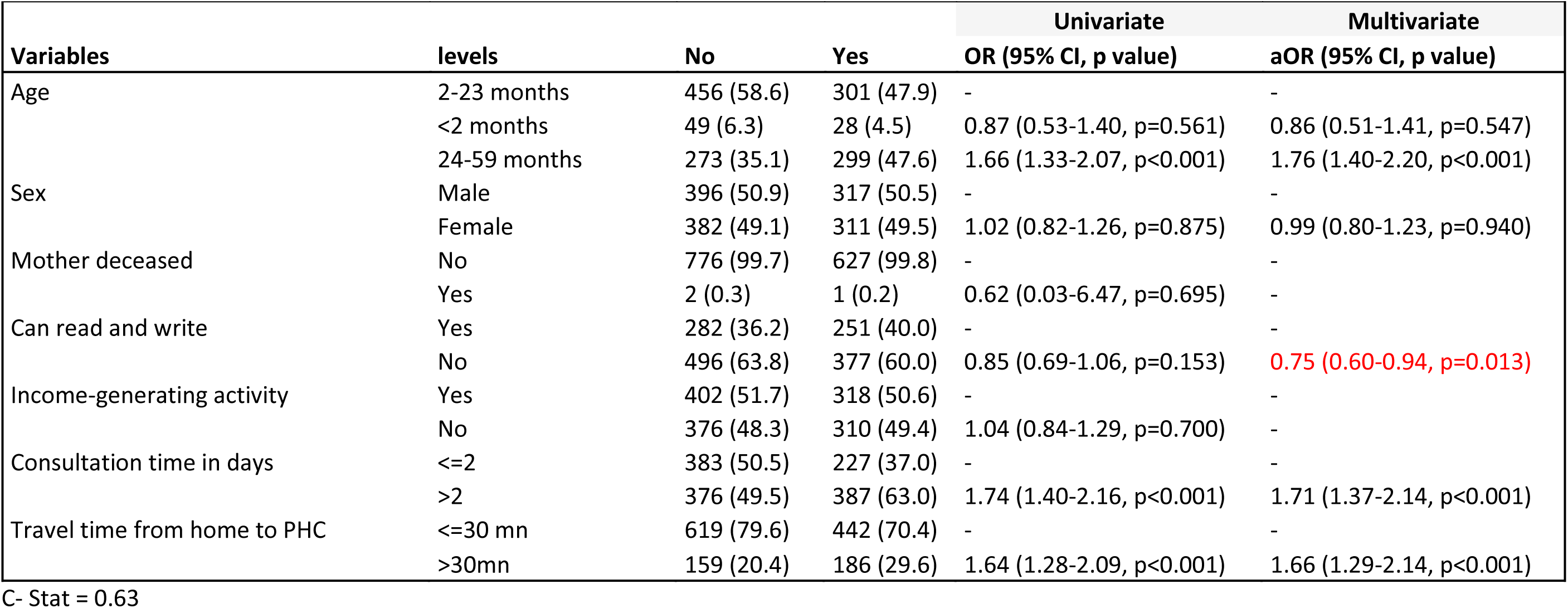
Factors associated to severity of cases identified at PHC in Guinea using a logistic regression model, AIRE research, June 2021 – June 2022 (n= 628 Vs 778)

| Variables | levels | No | Yes | Univariate | Multivariate |
| --- | --- | --- | --- | --- | --- |
|  |  |  |  | OR (95% CI, p value) | aOR (95% CI, p value) |
| Age | 2-23 months | 456 (58.6) | 301 (47.9) | - | - |
|  | <2 months | 49 (6.3) | 28 (4.5) | 0.87 (0.53-1.40, p=0.561) | 0.86 (0.51-1.41, p=0.547) |
|  | 24-59 months | 273 (35.1) | 299 (47.6) | 1.66 (1.33-2.07, p<0.001) | 1.76 (1.40-2.20, p<0.001) |
| Sex | Male | 396 (50.9) | 317 (50.5) | - | - |
|  | Female | 382 (49.1) | 311 (49.5) | 1.02 (0.82-1.26, p=0.875) | 0.99 (0.80-1.23, p=0.940) |
| Mother deceased | No | 776 (99.7) | 627 (99.8) | - | - |
|  | Yes | 2 (0.3) | 1 (0.2) | 0.62 (0.03-6.47, p=0.695) | - |
| Can read and write | Yes | 282 (36.2) | 251 (40.0) | - | - |
|  | No | 496 (63.8) | 377 (60.0) | 0.85 (0.69-1.06, p=0.153) | 0.75 (0.60-0.94, p=0.013) |
| Income-generating activity | Yes | 402 (51.7) | 318 (50.6) | - | - |
|  | No | 376 (48.3) | 310 (49.4) | 1.04 (0.84-1.29, p=0.700) | - |
| Consultation time in days | <=2 | 383 (50.5) | 227 (37.0) | - | - |
|  | >2 | 376 (49.5) | 387 (63.0) | 1.74 (1.40-2.16, p<0.001) | 1.71 (1.37-2.14, p<0.001) |
| Travel time from home to PHC | <=30 mn | 619 (79.6) | 442 (70.4) | - | - |
|  | >30mn | 159 (20.4) | 186 (29.6) | 1.64 (1.28-2.09, p<0.001) | 1.66 (1.29-2.14, p<0.001) |

C- Stat = 0.63

**Suplementary table3:**
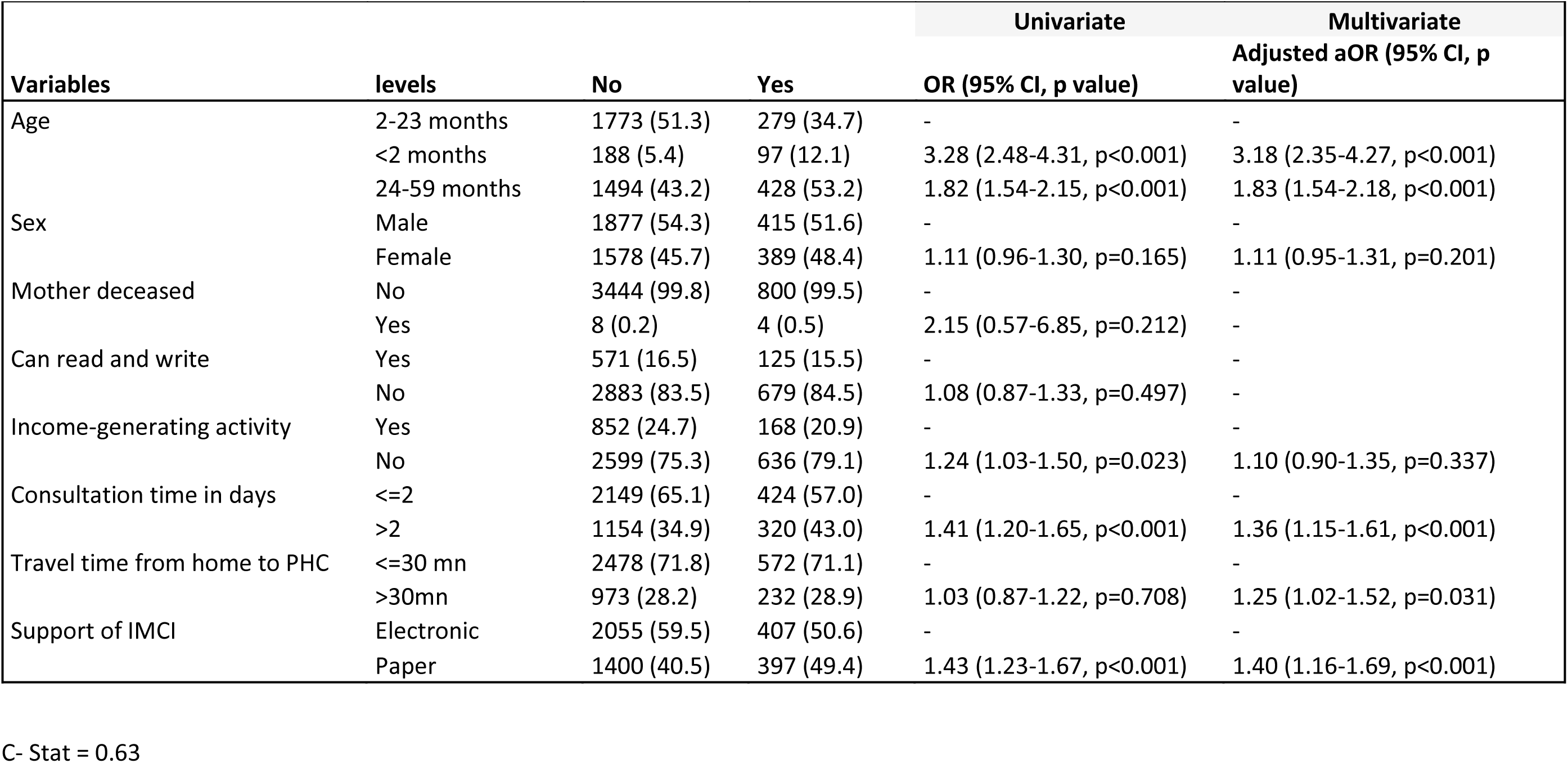
Factors associated to severity of cases identified at PHC in Mali using a logistic regression model, AIRE research, June 2021 – June 2022 (n= 804 Vs 3,455)

**Supplementary table 4:** Factors associated to severity of cases identified at PHC in Niger; AIRE research, June 2021 – June 2022 (n= 324 Vs 5,013)

| Logistic Regression Analysis: Niger |  |  |  |  |  |
| --- | --- | --- | --- | --- | --- |
| label | levels | No | Yes | Univariate<br>OR (95% CI, p value) | Multivariate<br>aOR (95% CI, p value) |
| Age | 2-23 months | 3089 (61.6) | 195 (60.2) | - | - |
|  | <2 months | 219 (4.4) | 41 (12.7) | 2.97 (2.04-4.23, p<0.001) | 1.97 (1.25-3.02, p=0.003) |
|  | 24-59 months | 1705 (34.0) | 88 (27.2) | 0.82 (0.63-1.06, p=0.127) | 0.86 (0.65-1.13, p=0.289) |
| Sex | Male | 2648 (52.8) | 169 (52.2) | - | - |
|  | Female | 2365 (47.2) | 155 (47.8) | 1.03 (0.82-1.29, p=0.817) | 0.99 (0.77-1.27, p=0.937) |
| Mother deceased | No | 4985 (99.5) | 319 (98.5) | - | - |
|  | Yes | 24 (0.5) | 5 (1.5) | 3.26 (1.09-7.92, p=0.017) | 1.88 (0.52-5.27, p=0.273) |
| Can read and write | Yes | 2368 (47.3) | 76 (23.5) | - | - |
|  | No | 2639 (52.7) | 248 (76.5) | 2.93 (2.26-3.83, p<0.001) | 2.43 (1.81-3.30, p<0.001) |
| Income-generating activity | Yes | 1070 (21.4) | 36 (11.1) | - | - |
|  | No | 3936 (78.6) | 288 (88.9) | 2.17 (1.55-3.15, p<0.001) | 1.41 (0.98-2.11, p=0.076) |
| Consultation time in days | <=2 | 3712 (78.4) | 120 (41.2) | - | - |
|  | >2 | 1023 (21.6) | 171 (58.8) | 5.17 (4.06-6.61, p<0.001) | 4.44 (3.46-5.71, p<0.001) |
| Travel time from home to PHC | <=30 mn | 4172 (83.3) | 218 (67.3) | - | - |
|  | >30mn | 837 (16.7) | 106 (32.7) | 2.42 (1.89-3.08, p<0.001) | 1.62 (1.23-2.11, p<0.001) |

C- Stat = 0.77

## Appendix

### ACKNOWLEDGMENTS

We thank all the children and their families who participated in the study, as well as the healthcare staff at the participating hospitals and the sites involved. We thank the field project staff and the AIRE Research Study Group*. We thank the Ministries of Health of the participating countries for their support.

#### Acknowledgements: Children, families, UNITAID and The AIRE Research Study Group

**Country investigators:** Ouagadougou, Burkina Faso: S. Yugbaré Ouédraogo (PI), V. M. Sanon Zombré (CoPI), Conakry, Guinea: M. Sama Cherif (CoPI), I. S. Diallo (CoPI), D. F. Kaba, (PI). Bamako, Mali: A. A. Diakité (PI), A. Sidibé, (CoPI). Niamey, Niger: H. Abarry Souleymane (CoPI), F. Tidjani Issagana Dikouma (PI). **Research coordinators & data centers: Inserm U1295, Toulouse 3 University, France:** H. Agbeci (Int Health Economist), L. Catala (Research associate), D. L. Dahourou (Research associate), S. Desmonde (Research associate), E. Gres (PhD Student), G. B. Hedible (Int research project manager), V. Leroy (research coordinator), L. Peters Bokol (Int clinical research monitor), J. Tavarez (Research project assistant), Z. Zair (Statistician, Data scientist). **CEPED, IRD, Paris, France:** S. Louart (process manager), V. Ridde (process coordination). **Inserm U1137, Paris, France :** A. Cousien (Research associate). **Inserm U1219,** EMR271 IRD, **Bordeaux University, France** : R. Becquet (Research associate), V. Briand (Research associate), V. Journot (Research associate). **PACCI, CHU Treichville, Abidjan, Côte d’Ivoire** : S. Lenaud (Int data manager), C. N’Chot (Research associate), B. Seri (Supervisor IT), C. Yao (data manager supervisor). **Consortium NGOs partners: Alima-HQ (consortium lead), Dakar, Sénégal**: G. Anago (Int Monitoring Evaluation Accountability And Learning Officer), D. Badiane (Supply chain manager), M. Kinda (Director), D. Neboua (Medical officer), P. S. Dia (Supply chain manager), S. Shepherd (referent NGO), N. di Mauro (Operations support officer), G. Noël (Knowledge broker), K. Nyoka (Communication and advocacy officer), W. Taokreo (Finance manager), O. B. Coulidiati Lompo (Finance manager), M. Vignon (Project Manager). **Alima, Conakry, Guinea:** P. Aba (clinical supervisor), N. Diallo (clinical supervisor), M. Ngaradoum (Medical Team Leader), S. Léno (data collector), A. T. Sow (data collector), A. Baldé (data collector), A. Soumah (data collector), B. Baldé (data collector), F. Bah (data collector), K. C. Millimouno (data collector), M. Haba (data collector), M. Bah (data collector), M. Soumah (data collector), M. Guilavogui (data collector), M. N. Sylla (data collector), S. Diallo (data collector), S. F. Dounfangadouno (data collector), T. I. Bah (data collector), S. Sani (data collector), C. Gnongoue (Monitoring Evaluation Accountability And Learning Officer), S. Gaye (Monitoring Evaluation Accountability And Learning Officer), J. P. Y. Guilavogui (Clinical Research Assistant), A. O. Touré (Country health economist), J. S. Kolié (Country clinical research monitor), A. S. Savadogo (country project manager). **Alima, Bamako, Mali:** F. Sangala (Medical Team Leader), M. Traore (Clinical supervisor), T. Konare (Clinical supervisor), A. Coulibaly (Country health economist), A. Keita (data collector), D. Diarra (data collector), H. Traoré (data collector), I. Sangaré (data collector), I. Koné (data collector), M. Traoré (data collector), S. Diarra (data collector), V. Opoue (Monitoring Evaluation Accountability And Learning Officer), F. K. Keita (medical coordinator), M. Dougabka (Clinical research assistant then Monitoring Evaluation Accountability And Learning Officer), B. Dembélé (data collector then Clinical research assistant), M. S. Doumbia (country health economist), G. D. Kargougou (country clinical research monitor), S. Keita (country project manager). **Solthis-HQ, Paris**: S. Bouille (NGO referent), S. Calmettes (NGO referent), F. Lamontagne (NGO referent). **Solthis, Niamey:** K. H. Harouna (clinical supervisor), B. Moutari (clinical supervisor), I. Issaka (clinical supervisor), S. O. Assoumane (clinical supervisor), S. Dioiri (Medical Team Leader), M. Sidi (data collector), K. Sani Alio (Country supply chain officer), S. Amina (data collector), R. Agbokou (Clinical research assistant), M. G. Hamidou (Clinical Research Assistant), S. M. Sani (Country health economist), A. Mahamane, Aboubacar Abdou (data collector), B. Ousmane (data collector), I Kabirou (data collector), I. Mahaman (data collector), I Mamoudou (data collector), M. Baguido (data collector), R. Abdoul (data collector), A. Sahabi (data collector), F. Seini (data collector), Z. Hamani (data collector), L-Y B Meda (Country clinical research monitor), Mactar Niome (country project manager), X. Toviho (Monitoring Evaluation Accountability And Learning Officer), I. Sanouna (Monitoring Evaluation Accountability And Learning Officer), P. Kouam (program officer). **Terre des hommes-HQ, Lausanne:** S. Busière (NGO referent), F. Triclin (NGO referent). **Terre des hommes, BF:** A. Hema (country project manager), M. Bayala (IeDA IT), L. Tapsoba (Monitoring Evaluation Accountability And Learning Officer), J. B. Yaro (Clinical reearch assistant), S. Sougue (Clinical reearch assistant), R. Bakyono (Country health economist), A. G. Sawadogo (Country clinical research monitor), A. Soumah (data collector), Y. A. Lompo (data collector), B. Malgoubri (data collector), F. Douamba (data collector), G. Sore (data collector), L. Wangraoua (data collector), S. Yamponi (data collector), S. I. Bayala (data collector), S. Tiegna (data collector), S. Kam (data collector), S. Yoda (data collector), M. Karantao (data collector), D. F. Barry (Clinical supervisor), O. Sanou (clinical supervisor), N. Nacoulma (Medical Team Leader), N. Semde (clinical supervisor), I. Ouattara (Clinical supervisor), F. Wango (clinical supervisor), Z. Gneissien (clinical supervisor), H. Congo (clinical supervisor). **Terre des hommes, Mali:** Y. Diarra (clinical supervisor), B. Ouattara (clinical supervisor), A. Maiga (data collector), F. Diabate (data collector), O. Goita (data collector), S. Gana (data collector), S. Diallo (data collector), S. Sylla (data collector), D. Coulibaly (Tdh project manager), N. Sakho (NGO referent). **Country SHS team: Burkina Faso:** K. Kadio (consultant and research associate), J. Yougbaré (data collector), D. Zongo (data collector), S. Tougouma (data collector), A. Dicko (data collector), Z. Nanema (data collector), I. Balima (data collector), A. Ouedraogo (data collector), A. Ouattara (data collector), S. E. Coulibaly (data collector). **Guinea**: H. Baldé (consultant and research associate), L. Barry (data collector), E. Duparc Haba (data collector). **Mali**: A. Coulibaly (consultant and research associate), T. Sidibe (data collector), Y. Sangare (data collector), B. Traore (data collector), Y. Diarra (data collector). **Niger**: A. E. Dagobi (consultant and research associate), S. Salifou (data collector), B. Gana Moustapha Chétima (data collector), I. H. Abdou (data collector)

### FOOTNOTES

#### Handling editor

##### Contributors

VL and VR conceptualised the research. The AIRE Research Study Group conducted training, data collection and management. ZZ with contributions from HGB and VL realise the data analysis. HGB prepared the first draft of this article. All authors were involved in data interpretation and review of the final manuscript. VL is the guarantor to submit the manuscript.

##### Funding

The AIRE project is funded by UNITAID, with in-kind support from Inserm and IRD. UNITAID was not involved in the design of the study, the collection, analysis and interpretation of the data, nor in the writing of the manuscript.

##### Competing interest

All authors have declared no conflict of interest.

##### Ethics approval and consent to participate

Ethics approval and consent to participate The AIRE research protocol, the information notice (translated in vernacular languages), the written consent form and any other relevant document have been submitted to each national ethics committee, to the Inserm Institutional Evaluation Ethics Committee (IEEC) and to the WHO Ethics Review Committee (WHO-ERC). All the aforementioned ethical committees reviewed and approved the protocol and other key documents (Comité d’Ethique pour la Recherche en Santé (CERS), Burkina Faso n°2020–4-070; Comité National d’Ethique pour la Recherche en Santé (CNERS), Guinea n°169/CNERS/21; Comité National d’Éthique pour la Santé et les Sciences de la vie (CNESS), Mali n°127/MSDS-CNESS; Comité National d’Ethique pour la Recherche en Santé (CNERS) Niger n°67/2020/CNERS; Inserm IEEC n°20–720; WHO-ERC n° ERC.0003364). This study has been retrospectively registered by the Pan African Clinical Trials Registry on June 15th 2022 under the following Trial registration number: PACTR202206525204526.

##### Data Availability Statement

The datasets generated and analysed during the current study are not publicly available. Access to processed deidentified participant data will be made available to any third Party after the publication of the main AIRE results stated in the Pan African Clinical Trial Registry Study statement (PACTR202206525204526, registered on 06/15/2022), upon a motivated request (concept sheet), and after the written consent of the AIRE research coordinator (Valeriane Leroy, Inserm U1295 Toulouse, France, orcid.org/0000-0003-3542-8616) obtained after the approval of the AIRE publication committee, if still active.

